# Cultural Adaptation and Linguistic Validation of a Community-Based Autism Screening Tools for Early Identification in Tanzania: A Mixed-Methods Study

**DOI:** 10.64898/2026.07.12.26357871

**Authors:** Alex Gabagambi Alexander, Catherine Mbughuni, Theonest Ndyetabura, Aloyce Mpepo, Rosalia Njau, Innocent Peter, Jovin Tibenderana, Thierry Kabwe, Elia Swai, Blandina Mmbaga, Florida Joseph Muro

**Author notes:** CORRESPONDING AUTHOR: (AG).

## Abstract

**Background:** In Tanzania and many other resource-limited settings, early diagnosis of Autism Spectrum Disorder (ASD) is often delayed due to a lack of culturally validated screening tools. Most available ASD screening tools were developed in Western countries and may not reflect local cultural and linguistic contexts.

**Purpose:** This study aimed to culturally adapt and linguistically validate the Swahili versions of both the Modified Checklist for Autism in Toddlers, Revised with Follow-Up (M-CHAT-R/F) and the Social Communication Questionnaire (SCQ) for use in the Tanzanian context.

**Methods:** A sequential mixed-methods study was conducted in Moshi Municipality and Hai District, Kilimanjaro Region, Tanzania. The study involved translation, cross-cultural adaptation, cognitive debriefing, expert panel review, and pilot testing of the psychometric evaluation of the M-CHAT-R/F and SCQ. Qualitative interviews and focus group discussions explored comprehension and contextual appropriateness of the tools. Pilot testing involved 32 caregivers of children aged 1–8 years. Internal consistency was assessed using Cronbach’s alpha and KR-20.

**Results:** Participants found both the adapted tools understandable and culturally appropriate. Unfamiliar examples, such as vacuum cleaners and dinosaurs, were replaced with locally familiar examples including domestic animals, loud music, and vehicle sounds. The M-CHAT-R/F demonstrated acceptable internal consistency (Cronbach’s α = 0.77; KR-20 = 0.79), while the SCQ showed good reliability (Cronbach’s α = 0.83; KR-20 = 0.84). Cognitive debriefing and expert review confirmed the clarity and comprehension of the adapted tools for community use.

**Conclusion:** The adapted Swahili versions of the M-CHAT-R/F and SCQ demonstrated promising preliminary reliability and cultural relevance for ASD screening in Tanzania. These findings support the use of culturally tailored screening tools to improve early identification and referral of children with ASD in Tanzania.

## Introduction

Autism spectrum disorder (ASD) is a neurodevelopmental condition characterized by persistent challenges in social communication and social interaction, as well as restricted and repetitive behaviors and interests (Abubakar et al., 2016; Hodges et al., 2020). Worldwide, an estimated 61.8 million people live with ASD, equivalent to approximately 1 in 127 people (Santomauro et al., 2024). The prevalence of ASD has continued to increase in high-income countries as well as Low- and Middle-Income Countries (LMICs) (Hine et al., 2020). In Africa, the prevalence of ASD is estimated to be around 1%, although the true extent of the problem is likely to be higher due to limited screening services and lack of early detection programs, particularly in Sub-Saharan Africa (SSA) (Salari et al., 2022; Sobieski et al., 2022). In Tanzania and other SSA countries, early diagnosis of ASD is still delayed, underscoring the importance of having accessible, culturally sensitive, and community-based screening methods (Ruparelia, 2021).

Early identification of ASD is critical, as timely intervention is associated with improved developmental outcomes, including better communication, social functioning, and adaptive skills (Nourzad, n.d.; Salari et al., 2022; Snijder et al., 2022). Although ASD can often be detected within the first two years of life, early diagnosis remains challenging in LMICs due to limited awareness, absence of culturally adapted tools, inadequate specialist services, and constrained healthcare infrastructure (Harrison et al., 2016; Janvier et al., 2016). Heavy reliance on clinical professionals further contributes to delayed diagnosis, particularly in resource-limited settings where access to specialized care is scarce (Harrison et al., 2016; Khowaja et al., 2015). These delays reduce opportunities for early intervention and contribute to long-term developmental inequities (Janvier et al., 2016). Addressing these barriers therefore requires accessible strategies that can identify children at risk before delays become more severe. Community-based early detection approaches offer a promising solution to these challenges. Screening initiatives implemented in non-clinical settings, such as preschools and daycare centers, have been shown to improve early identification and referral rates (Gulsrud et al., 2019). These environments often represent children’s first structured social interactions outside the home, providing valuable opportunities to observe developmental patterns (Janvier et al., 2016). Early childhood caregivers and teachers interact with children daily and are well-positioned to identify atypical behaviors by comparing them with peers (D. Zhang et al., 2019). However, their role in ASD identification remains underutilized due to the lack of simple, validated screening tools suitable for community use (Brewer, 2020). Strengthening these platforms through culturally appropriate tools could significantly enhance early detection in underserved settings (Janvier et al., 2016).

Several screening tools have been developed for early ASD detection, including the Autism Parent Screening Instrument (APSI), Brief Infant-Toddler Social and Emotional Assessment (BITSEA), Checklist for Early Signs of Developmental Disorders (CESDD), Communication and Symbolic Behavior Scales Developmental Profile (CSBS-DP), Modified Checklist for Autism in Toddlers (M-CHAT), Social Attention and Communication Surveillance (SACS), Social Communication Questionnaire (SCQ), and Screening Tool for Autism in Toddlers and Young Children (STAT) (Han et al., 2023; Janvier et al., 2016; Sobieski et al., 2022; Y. Zhang et al., 2022). Among these, M-CHAT and SCQ are widely used in community settings (Han et al., 2023; Sobieski et al., 2022; Y. Zhang et al., 2022). The M-CHAT-R/F demonstrates strong performance, with a pooled sensitivity of 0.83 and a specificity of 0.94, particularly when combined with follow-up interviews (Wieckowski et al., 2023), while the SCQ shows moderate diagnostic accuracy (Sangare et al., 2019; Staton et al., 2023). However, most tools were developed in high-income contexts and may not adequately capture cultural and linguistic differences in LMICs, including Tanzania (Sobieski et al., 2022). For instance, items assessing eye contact, play, or gestures may not align with local child-rearing practices, leading to misinterpretation and reduced accuracy (Ezmeci et al., 2022). Therefore, cultural adaptation is essential to ensure the validity and applicability of ASD screening tools. This process involves not only translation, but also contextual modification based on local norms and developmental expectations. Studies in African settings highlight this need: in South Africa, the Northern Sotho adaptation of M-CHAT required substantial modification to ensure cultural relevance (Vorster et al., 2021); in Mali, validation studies emphasized adapting items to local understandings of child behavior (Sangare et al., 2019); and in Uganda, a locally adapted screening tool demonstrated good sensitivity and specificity for developmental disabilities, including ASD (Kakooza-Mwesige et al., 2014). These findings underscore the importance of culturally grounded validation processes.

Although these tools have been adapted in several LMICs, there remains limited evidence on culturally validated Swahili-language tools suitable for community-based use in Tanzania(Harrison et al., 2014). Although some efforts have been made to adapt clinical instruments such as the Childhood Autism Rating Scale (CARS2), their use in community settings remains limited, as they are designed for trained clinicians and are not readily applicable by lay community members with limited training (Harrison et al., 2014; Huerta & Lord, 2012). Additionally, linguistic diversity and regional variations in child development norms further complicate the use of standardized tools (Leighton et al., 2023). These challenges are particularly evident in underserved regions, where access to pediatric diagnostic services is limited and early detection opportunities are often missed.

This study aimed to culturally adapt, linguistically validate, and preliminarily evaluate the psychometric properties of a simple, community-based ASD screening tool for use in Tanzania through participatory engagement with caregivers, educators, and health workers. By leveraging community-based platforms and strengthening early detection capacity, this work aims to support scalable strategies for improving early ASD identification and developmental outcomes in resource-limited settings.

## Methodology

### Study Design

We employed a sequential exploratory mixed-methods study design to translate, culturally adapt, linguistically validate, and preliminarily evaluate the psychometric properties of ASD screening tools for use. The study consisted of four interconnected phases: (1) tool selection and preparatory stakeholder engagement, (2) translation and cross-cultural adaptation, (3) linguistic validation and cognitive pretesting, and (4) pilot psychometric evaluation. The approach was guided by the Social Ecological Model (SEM), and engagement of different stakeholders was used to culturally adapt and linguistically validate the M-CHAT-R/F and SCQ screening tools for use in Moshi and Hai districts, Tanzania (McLeroy et al., 1988; Stokols, 1996).

### The Conceptual Framework

The SEM considers multiple contextual dimensions, namely, the micro-level, meso-level, and macro-level, to understand and address the complex factors influencing the cultural adaptation process of the ASD tool in the Kilimanjaro region of Tanzania (Graif et al., 2021; McLeroy et al., 1988; Stokols, 1996). The SEM is a theoretical framework that is used to examine how individual behavior and health outcomes are influenced not only by personal factors but also by a broader network of social, environmental, and policy-related influences (Graif et al., 2021). Applying SEM to this study allows for holistic implementation of an ASD screening tool at multiple, interconnected levels (figure1). This approach guided the adaptation of the screening tool to ensure it is culturally relevant, contextually appropriate, and feasible for use across different levels of the community. It will also inform the development of multi-tiered strategies for training, community engagement, and advocacy aimed at improving early ASD detection and long-term outcomes for children in the study area.

**Figure 1:**
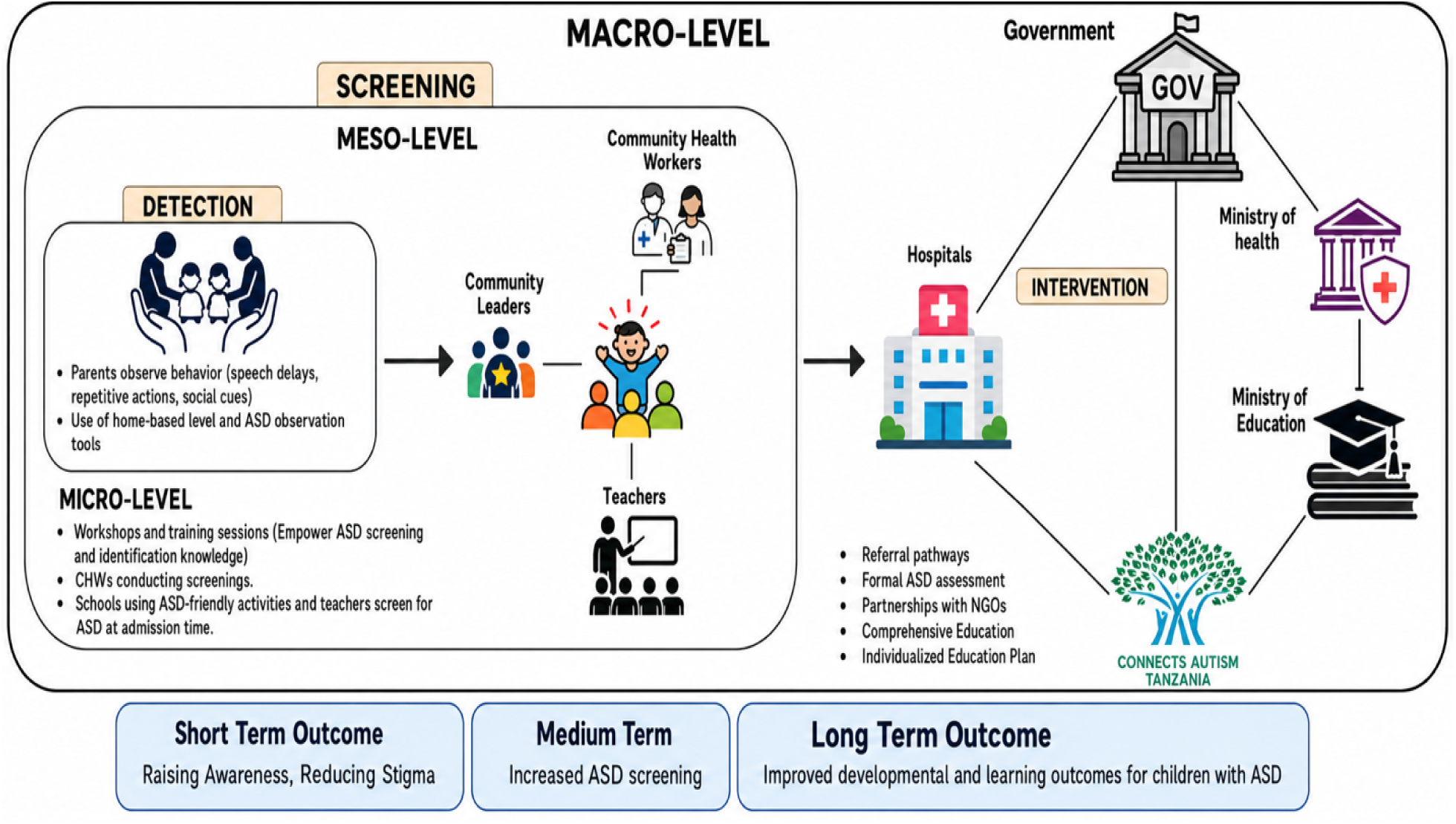
The Modified Social Ecological Model

### Study Setting and Sampling

The study was conducted in selected preschools in Moshi Municipality and Hai District in the Kilimanjaro Region of northern Tanzania, including Moshi, Shirimatunda, Kongoroni, Gabriella Rehabilitation Centers, Juhudi, Faraja Primary, Mabogini, and Mwereni. These settings represent urban and peri-urban communities. Data were collected within preschool environments. A two-stage sampling approach was used: snowball sampling to identify schools with children with ASD, followed by purposive sampling to recruit preschool caregivers and parents as participants.

### Translation, Cross-Cultural Adaptation, and Linguistic Validation of ASD Screening Tool

#### Study Design

This phase employed a qualitative design to explore clarity, comprehension, and cultural relevance of the M-CHAT-R and the SCQ during translation and adaptation for use in Tanzania.

#### Study Population and Stakeholder Engagement

The study engaged stakeholders across the SEM, including caregivers of children aged 1–8 years, preschool teachers, daycare providers, administrators, clinic managers, community and religious leaders, NGOs, policymakers, and a multidisciplinary expert panel. Caregivers provided insights on child development and help-seeking behaviors, educators contributed classroom observations, organizational stakeholders informed feasibility, community actors addressed cultural beliefs and stigma, and policymakers guided system integration.

#### Inclusion and Exclusion Criteria

Participants were required to be fluent in Swahili (or English for policymakers/experts), reside or work in Moshi and Hai districts, and provide informed consent. Caregivers were the primary caregivers of children aged 1–8 years. Educators and health providers are required to have at least six months of experience. NGO representatives held relevant leadership roles, while experts had recognized disciplinary expertise. Individuals unwilling to consent, temporary residents, or those with significant cognitive impairment were excluded.

#### Tool Selection

The study adapted the M-CHAT-R and SCQ; these instruments were selected due to strong psychometric properties (Dai et al., 2021; Perera et al., 2017), they are appropriate for the age (Dai et al., 2021; Marvin et al., 2017; Nwokolo et al., 2024a; Oner & Munir, 2020a), and they are feasible (Dai et al., 2021; Perera et al., 2017). The SCQ requires 10-15 minutes (Kipkemoi et al., 2025a; Marvin et al., 2017). The M-CHAT-R/F screens toddlers aged 16–30 months, while the SCQ targets children aged 4 years and older, enabling developmental coverage across early childhood.

#### Translation, Cross-Cultural Adaptation Process, and Stakeholder Engagement

Translation and cross-cultural adaptation of the M-CHAT-R/F and the SCQ were conducted following the guidelines of the International Society for Pharmacoeconomics and Outcomes Research (ISPOR) to ensure linguistic accuracy, cultural relevance, and conceptual equivalence between the original and translated versions. The process began with forward translation, where each instrument was translated from English to Swahili by two independent translators with proficiency in both languages. Later, the translations were compared and merged to obtain a single agreed version (reconciliation). This version was back-translated from Swahili to English by two other independent translators who had not seen the original versions of the instruments.

Subsequently, a multidisciplinary expert panel reviewed all versions to assess conceptual equivalence, face validity, cultural appropriateness, linguistic accuracy, and feasibility using specific evaluation forms and a modified Delphi consensus process (Boulkedid et al., 2011; Nasa et al., 2021). Pre-final versions of both instruments were evaluated through cognitive debriefing interviews, which involved at least two participants from each stakeholder group, including caregivers, teachers, school principals, and key stakeholders. These interviews aimed to assess the level of comprehension, clarity, acceptability, and cultural relevance of each questionnaire item. Feedback from cognitive debriefing was used to make iterative revisions, which led to the final Swahili versions of the M-CHAT-R/F and SCQ.

Guided by the SEM, 10 IDIs and 3 FGDs were conducted across all stakeholder levels. Cognitive interviews assessed comprehension, acceptability, and cultural relevance. Key adaptations included replacing unfamiliar idioms with local examples, aligning items with local developmental norms, and ensuring yes/no response formats suited community contexts. A diverse panel of 10 participants was recruited to ensure representation across urban and rural contexts, gender balance, and professional diversity, mirroring inclusive stakeholder engagement models used in similar global health studies(Coulter et al., 2019; Jackson et al., 2025). An interdisciplinary expert panel then evaluated each translated item using structured forms assessing face validity, content relevance, cultural sensitivity, language accuracy, and operational feasibility(Boulkedid et al., 2011). A panel then evaluated each translated item using structured forms assessing face validity, content relevance, cultural sensitivity, language accuracy, and operational feasibility(Boulkedid et al., 2011). Participants rated items on a five-point Likert scale (1 = agree, 2 = somewhat agree, 3 = unsure, 4 = somewhat disagree, 5 = disagree), with consensus defined as >70% scoring 1 or 2 and <25% scoring 3, 4, or 5 (Boulkedid et al., 2011; Nasa et al., 2021).

#### Pilot Testing and Cognitive Debriefing

A pilot study involving 32 caregivers assessed some psychometric properties, usability, and acceptability. Cognitive debriefing explored understanding, interpretation, recall processes, and cultural relevance. Feedback informed iterative refinement of items. Children were not directly involved; caregivers responded.

##### Data Collection and Analysis

We analyzed qualitative data by using a hybrid rapid thematic analysis approach, supported by NVivo version 15. The “hybrid” approach combined deductive coding, informed by the study objectives and interview guide. Inductive coding was conducted to allow new themes to emerge from the data. Initially, one team member reviewed each transcript, generated summaries, and conducted preliminary coding within NVivo. A second team member independently reviewed the coded transcripts to ensure accuracy, consistency, and coherence. Discrepancies were resolved through team discussions, and the coding framework was refined accordingly. Analytical memos were developed and cross-analyzed to identify overarching themes and subthemes. In addition, the quantitative data included descriptive statistics and psychometric testing. Internal consistency was assessed using Cronbach’s alpha and KR-20, with item-total correlations and “alpha if item deleted” examined. No items were removed, consistent with standardized tool integrity.

After separate quantitative and qualitative analyses, findings were integrated through methodological triangulation to provide a comprehensive understanding of comprehension, acceptability, and cultural relevance of both tools.

#### Ethical Considerations

Ethical approval for the study was sought from the KCMC University Research Ethics Review Committee (KURERC) of KCMC University (certificate No. 2789) and the National Health Research Ethics Review Committee (NatHREC) (certificate No. NIMR/HQ/R.8a/Vol.IX/5215). Written informed consent was obtained from all participants (parents or legal guardians), ensuring participation is voluntary and without coercion. Participants were informed that they can withdraw at any time without consequences. Parents and caregivers were fully informed about the study objectives, procedures, potential risks, and benefits through information sheets and, where appropriate, group or individual meetings. This approach ensures that caregivers are active partners in the decision-making process and can support the child throughout participation.

## Results

### Socio-demographic Characteristics of IDI Participants

The study respondents were predominantly female (90%), with a median age of 33 years (IQR: 28–39). The majority of participants had at least a diploma or bachelor’s degree in education, and represented a variety of roles, including health and allied health professionals, special education teachers, caregivers, NGO workers, and leaders of institutions and rehabilitation centers. The majority of participants (80%) reported having received training on child development and autism or developmental delay. Awareness of ASD was high (90%), although the primary source of information about autism varied, with health centers and schools being cited most frequently, along with other informal or informal sources (Table 1).

**Table 1.**
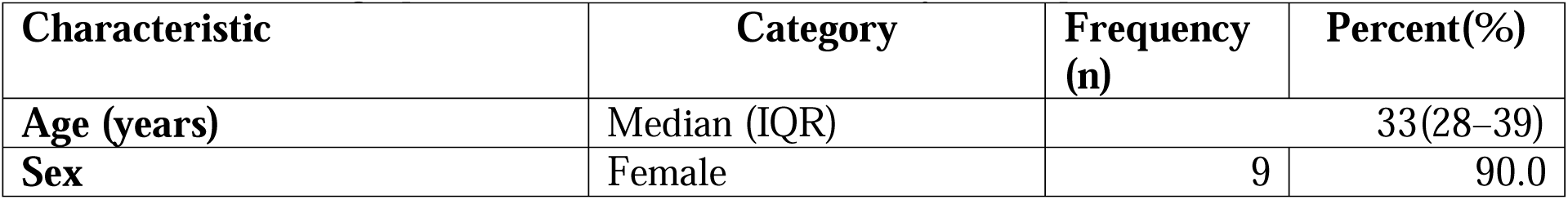
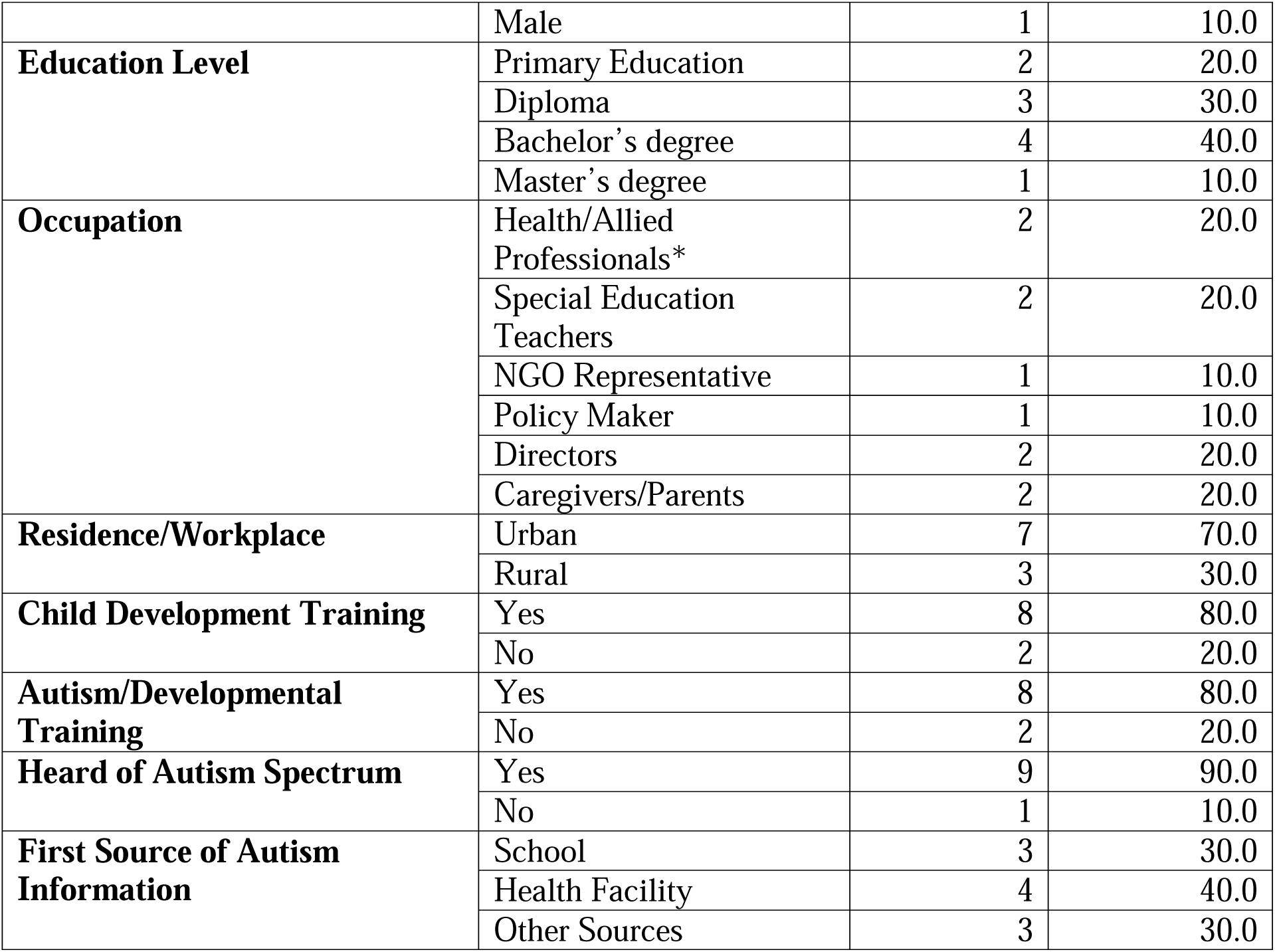
Sociodemographic Characteristics of IDI Study Participants (N = 10)

#### The sociodemographic characteristics of FGD participants

The sociodemographic characteristics of FGD participants, including occupational therapists, parents/caregivers, and teachers. Participants varied in age, education, work setting, and autism-related experience, providing diverse stakeholder perspectives. Females predominated across groups, particularly among teachers. Occupational therapists reported the highest levels of child development and autism training, while training among caregivers and teachers was limited.

**Table 2.**
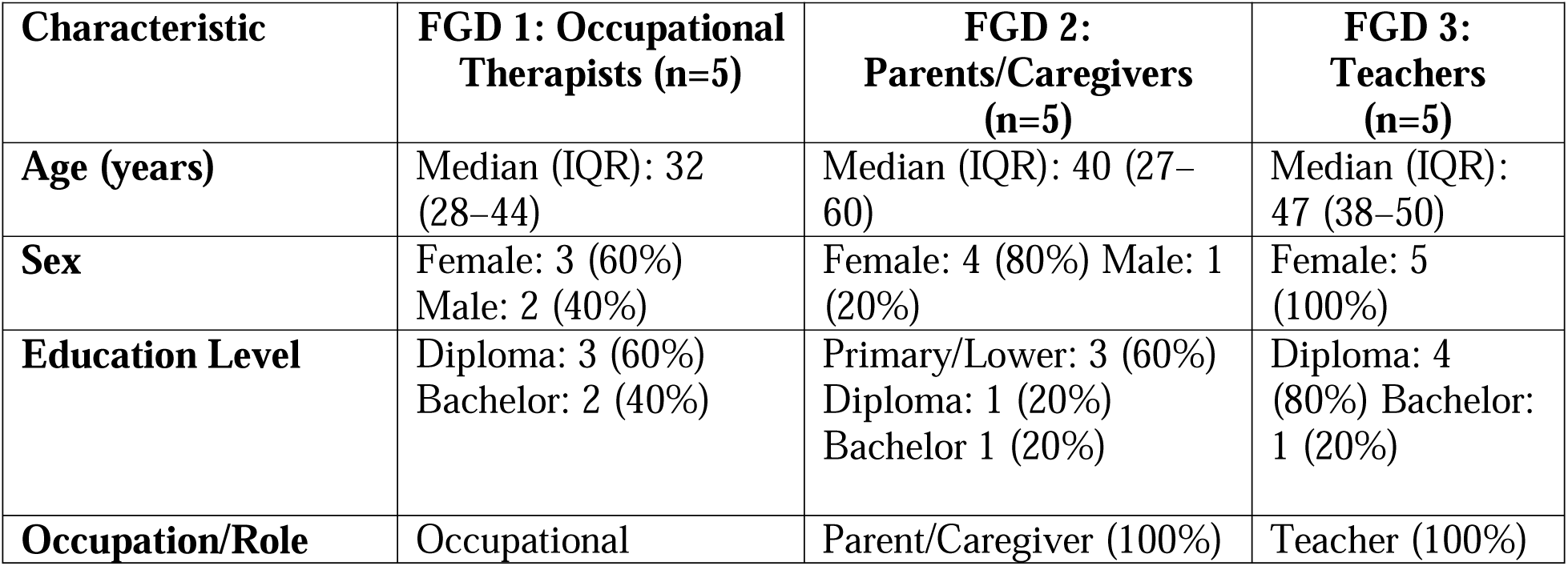
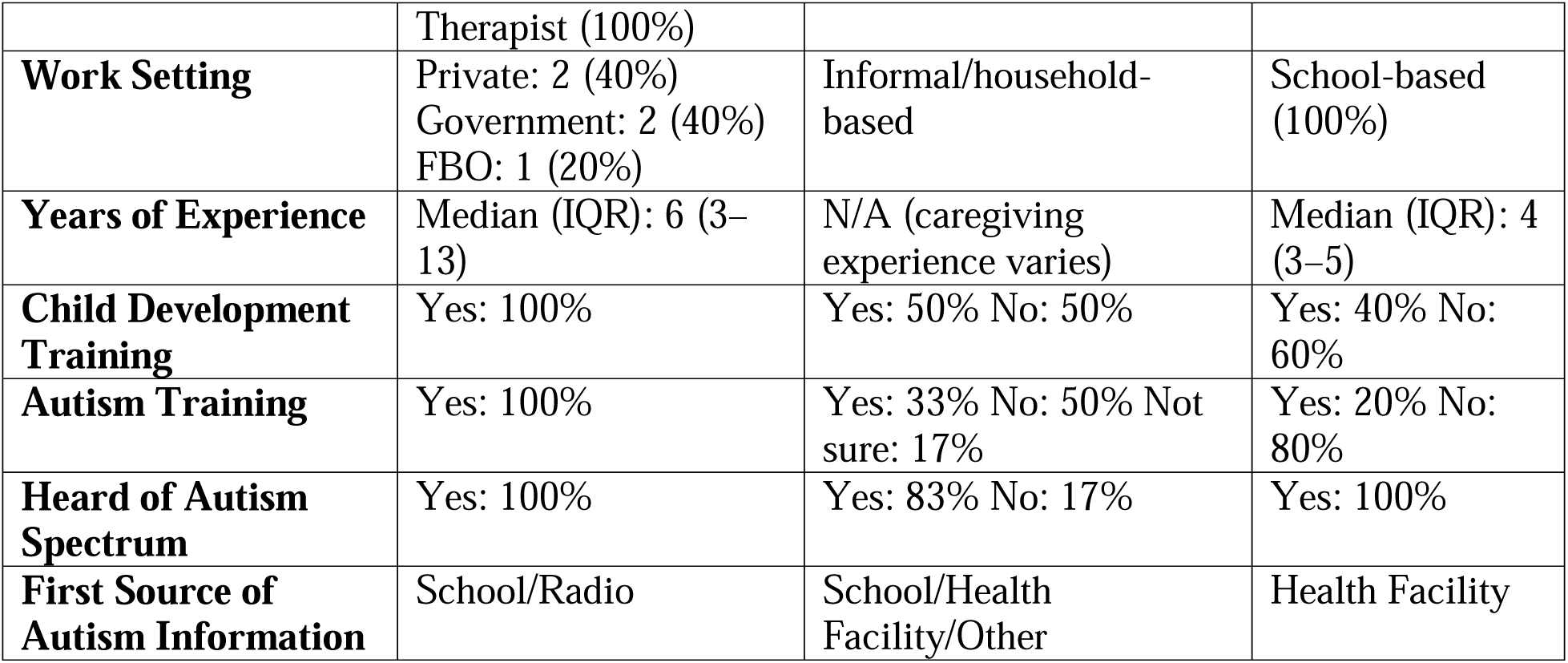
Sociodemographic Characteristics of FGDs Study Participants (N = 15)

### Translation, Cultural Adaptation, and Cognitive Debriefing

Feedback from these cultural adaptation and cognitive debriefing interviews informed additional refinements, particularly for items referencing uncommon objects or behaviors, as well as for phrasing that could obscure intended meaning.

Cognitive debriefing revealed that the questionnaires were generally understandable and self-explanatory.

> *“I have seen the questions, and they are understandable. However, I feel that there are some things we might need to understand more deeply.”* R1, Caregivers, FGD2)
>
> *“I’ve noticed that the way these questions are written, they are self-explanatory, so a person knows what to do or how to answer.”* (R5, Teacher, FGD1).

Some participants noted that the items mirrored questions typically asked in local child assessments and clearly reflected child developmental behaviors.

> *“Yes, these types of questions are commonly used here in our center. Many of the questions are similar to those we ask parents or guardians when assessing children.”* (Rehabilitation center’s (Director, IDI4)

Some participants highlighted occasional challenges with Swahili phrasing and the binary “Yes/No” response format, which could inadequately capture intermittent behaviors. They recommended incorporating a middle response category using a Likert-type scale to better reflect behaviors that occur occasionally or vary across contexts.

> *“Because what happens is that some children do things sometimes, others often, others most of the time, and others never. If we only have Yes and No, we will miss some information when we conclude. We might score that the child has no problem, while in reality, there are mild issues that could have been identified and addressed. With the current format, someone might finish the questionnaire and conclude the child has no problem, while some important details were missed.”* (Rehabilitation center’s Director, IDI3).

Conceptual clarity was another key theme: Stakeholders emphasized that ASD should be framed as a developmental condition rather than a disease. Professional respondents noted that most items were straightforward, though some terms required clarification for broader recognition.

> *“My question is this: Is autism a disease or a condition? Because, from our understanding, we consider it a condition, not a disease […] Most of the time we refer to it as a condition.”* (Special Needs Teacher, IDI1)

Cultural relevance emerged as a critical consideration. References to unfamiliar objects, such as vacuum cleaners or dinosaurs, were replaced with locally recognizable examples like domestic animals (cows, goats, elephants) and common environmental stimuli, including loud music or vehicle sounds. Similarly, play-based items were adapted to reflect materials accessible in local settings, such as sticks, rocks, or pencils, rather than standardized building blocks.

> *“Another issue is question twelve, which mentions a vacuum cleaner. This does not fit well in our context. Many families, especially in rural areas, do not use vacuum cleaners. Mothers might not understand what a machine that sucks dust is. Even when I read the question, I had to pause and think about it […] You could replace it with something more familiar, such as loud music or loud sounds. These are more common and easily understood.”* (Rehabilitation center’s Director, IDI4).
>
> *“We could use animals that people commonly know, such as a lion, leopard, or elephant”* (R4, Caregiver, FGD2)

Across all groups, participants confirmed that the tools reflected behaviors commonly observed in children with autism, and minor refinements were sufficient to enhance clarity and usability.

> *“They reflect behaviors and developmental milestones commonly observed in children.”* (NGO representative, IDI6).

#### Expert Panel Review

The expert review panel included 10 participants with a median age of 33 years (IQR: 28–46) and a balanced sex distribution. Members were well qualified, holding bachelor’s, master’s, and diploma-level education, and represented diverse roles including occupational therapists, physicians, policy makers, physiotherapists, psychiatrists, and administrators. Most were urban-based (90%) with a median of 5 years’ experience (IQR: 3–12). Nearly all had training in child development and autism (90%), providing multidisciplinary expertise for tool validation. An expert panel review was used to evaluate the relevance, clarity, and cultural appropriateness of the adapted items. Before the expert panel reviewed the Delphi ratings of the tools, which ranged from 3.0 to 5.0 (mean = 3.94), indicating general agreement on item suitability and the need for only minor contextual and linguistic adjustments (Table D1).

**Table 3:** Sociodemographic Characteristics of Expert Panel Review Participants (N = 10)

### Cultural Adaptation of the SCQ and M-CHAT During Expert Panel Review

Initially, face validity of the SCQ and the M-CHAT was assessed through expert review and stakeholder feedback during the pilot and adaptation process. Overall, participants reported that the tools were relevant and appropriate for assessing early social communication behaviors and developmental concerns in children.

The expert panel feedback in the initial round highlighted some challenges with linguistic clarity and comprehension, cultural relevance, and contextual appropriateness of the tools.

Under the theme of clarity and comprehension of language, some experts expressed concerns regarding the use of difficult Swahili terminology that could limit understanding among caregivers. One participant noted,

> *“Some Swahili words used here are a bit difficult. Even as a professional, I find some words challenging to understand. So, what about a parent, won’t they also struggle?”* (R6).

This highlighted the need for simplified and widely understood language in the adapted tools. The theme of Contextual Relevance reflected the importance of using culturally familiar examples to improve the interpretation of items. Participants recommended incorporating examples commonly encountered in local households and communities. As one reviewer explained,

> *“We should use things commonly found in households. For example, a blender or the sound of a refrigerator. These are more relatable”* (R4). Another expert added, *“For animals, we should include common animals”* (R10), emphasizing the importance of contextual familiarity in item comprehension.

In the theme item adaptation and translation, some reviewers suggested modifications to terminology and behavioral descriptions to improve clarity. For instance, one participant stated,

> *“In question 12 (MCHAT), it says ‘is more interested in a mwanasesere (doll).’ Not everyone understands the word ‘mwanasesere’. Why don’t we just use the word ‘mdoli’ or both of them?”* (R6).

Another reviewer proposed using clearer behavioral terminology, explaining,

> *“Question 15(MCHAT): ‘Does your child try to do things that you do?’ I suggest we use the word ‘imitate’”* (R5).

Revisions of both tools followed the expert panel review during the workshop, final expert ratings increased, and they confirmed great improvement with new final scores ranging from 4.5 to 5.0 (median 5.0). Slightly lower scores (4.5–4.7) were attributed to the length of the SCQ, with 40 questions; the time required for its administration in community settings was reported to be a challenge. They insisted on the need for comprehensive training for community rehabilitation staff and teachers in using both tools. Together, the translation and adaptation process, cognitive debriefing, Delphi consensus, and SEM-informed stakeholder engagement established that the Swahili versions of the M-CHAT-R and SCQ were understandable, culturally appropriate, and highly relevant for both caregivers and professionals. Minor adjustments informed by iterative feedback enhanced clarity, ensured conceptual accuracy, and supported local implementation across diverse urban and rural settings.

#### Pilot testing of MCHAT-R and SCQ

##### Study Participant Characteristics

A total of 32 caregivers participated in the pilot testing of the translated screening tools across eight early childhood sites in Moshi and Hai districts. The M-CHAT-R/F was administered to caregivers of children aged <4 years (n = 17), while the SCQ was administered to caregivers of children aged ≥4 years (n = 15). The sample comprised 29 males and 17 females. The median completion time was 7 minutes for the M-CHAT-R/F and 12 minutes for the SCQ.

##### Psychometric Properties of the M-CHAT (Less than 4 years)

The internal consistency of the 20-item M-CHAT R was examined in a sample of 17 respondents using both Cronbach’s alpha (α = 0.77) and Kuder–Richardson 20 (KR-20 = 0.79). Reverse-coded items (Q5, Q11, Q20) were recoded before the conduct of our analysis. Examination of item–total correlations indicated that several items (Q1, Q3, Q5, Q6, Q9, Q13, Q14, Q15) contributed strongly to the scale’s internal consistency (r > 0.50), while others performed moderately (Q4, Q7, Q8, r = 0.30–0.49). A few items exhibited weak or negative correlations (Q11, Q12, Q17, Q18, Q19). Deleting Q11 or Q19 marginally increased reliability (α ≈ 0.79), but no items were removed, consistent with the standard structure of the instrument (Figure 2).

**Figure 2:**
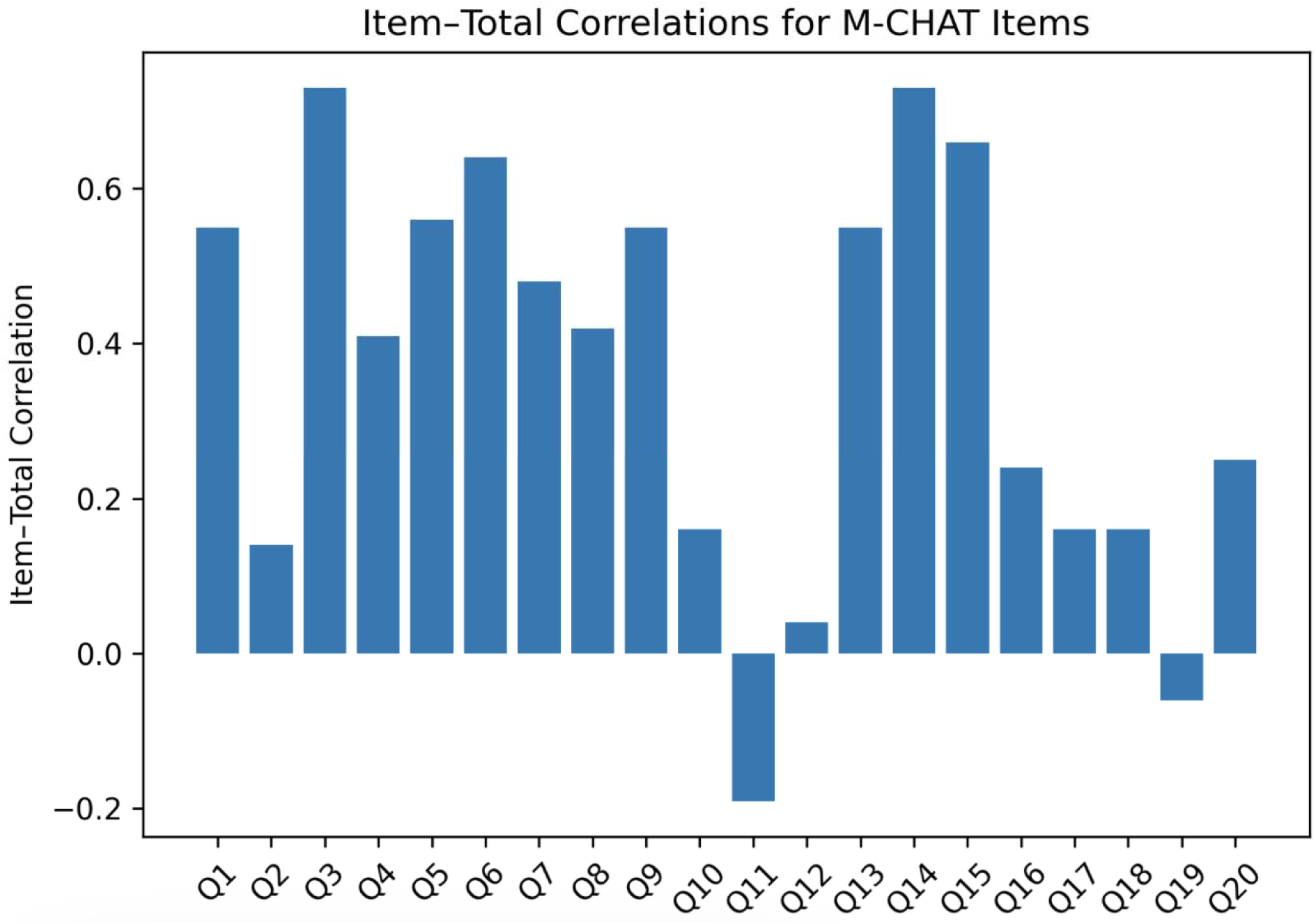
Conceptual structure of the M-CHAT screening instrument showing individual items contributing to the total score.

##### Psychometric Properties of the SCQ (Above 4 years)

The internal consistency of the SCQ was evaluated using Cronbach’s alpha and the Kuder–Richardson Formula 20 (KR-20), which is appropriate for dichotomously scored items. Prior to analysis, reverse-scored items (Q2, Q9, Q19, Q23, Q28, Q29, and Q33) were recoded to ensure consistent scoring direction. The overall scale demonstrated good internal consistency in the present sample, with a Cronbach’s α of 0.83 and a KR-20 of 0.84.

Item–total correlations ranged from −0.47 to 0.68, with most items demonstrating moderate to strong correlations with the total score. A small number of items, particularly those that were reverse-coded, showed weaker or negative item–total correlations. Examination of “Cronbach’s alpha if item deleted” indicated that removal of any single item would not substantially improve the overall reliability of the scale, as alpha values remained within a narrow range (approximately 0.82–0.85).

## Discussion

The study participants from the cognitive debriefing and expert panel review interviews revealed that there is a need to adapt both questionnaire items to ensure cultural relevance and contextual familiarity. Participants of this study recommended replacing unfamiliar references or examples, such as the use of vacuum cleaners or dinosaurs, with locally relevant examples, including domestic animals (e.g., cows, goats, and elephants) and common environmental stimuli such as loud music or vehicle sounds. This suggests that culturally contextualized examples are essential to ensure that caregivers accurately interpret screening questions. These findings are consistent with evidence from cross-cultural research on autism screening tools, which indicates that translation alone is insufficient for effective use in diverse settings. Instead, screening instruments often require cultural adaptation to ensure conceptual and contextual equivalence in the target population(Huda et al., 2024). For example, several reviews of autism screening tools in culturally and linguistically diverse populations reported that modifications to language, examples, instrument content, and contextual references are frequently necessary to maintain the validity of instruments across different cultural settings and everyday experiences (Al Maskari et al., 2018; Huda et al., 2024; Soto et al., 2015). In the study by Huda et al(2024), it was revealed that the screening tools developed in Western contexts may include examples or behavioral references that are unfamiliar in other cultures, which can affect caregivers’ interpretation and the accuracy of responses (Huda et al., 2024).

The 20-item M-CHAT-R demonstrated acceptable internal consistency, with Cronbach’s alpha of 0.77. These values suggest a reasonable level of consistency in measuring early autism-related social communication behaviors. Several items contributed strongly to the overall scale reliability, while a small number showed weaker or negative item–total correlations. These findings are consistent with previous studies examining the reliability of the M-CHAT-R in different populations by Robins et al (2014), who reported acceptable internal consistency for the tool in large validation samples, supporting its reliability as a screening instrument for ASD (Robins et al., 2014). Similarly, other validation studies conducted in diverse cultural settings have reported Cronbach’s alpha values ranging from approximately 0.70 to 0.90, indicating moderate to good reliability of the tool(Nukeshtayeva et al., 2022; Oner & Munir, 2020b; Tsai et al., 2019). These results suggest that the M-CHAT-R maintains stable psychometric properties across contexts. SCQ demonstrated good internal consistency, with Cronbach’s α of 0.83. This result reveals that the items of the scale were generally consistent in measuring behaviors associated with ASD. Although most items showed moderate to strong item–total correlations, a few, particularly reverse-coded items, displayed weaker or negative correlations. However, removal of any individual item did not substantially improve the reliability of the scale, suggesting that the overall structure of the instrument remained stable in the study sample. These findings are consistent with earlier psychometric evaluations of the SCQ. For example, studies validating the SCQ in different cultural settings have reported Cronbach’s alpha values ranging from approximately 0.80 to 0.90, indicating good reliability across populations(Kipkemoi et al., 2025b; Nwokolo et al., 2024b). The presence of lower correlations among some reverse-scored items has also been documented in previous research and may reflect differences in how respondents interpret negatively worded questions (İLhan et al., 2024).

## Study Limitations

This study faced several limitations, such as recruitment challenges, which may have risen due to stigma surrounding ASD, addressed through collaboration with local leaders, NGOs, peer recruiters, and participant compensation. Cultural adaptation limitations were mitigated through forward–backward translation, iterative piloting, FGDs, and validation with clinical assessment. Implementation constraints were managed using brief tools, mobile-based data collection, and established referral networks. Ethical risks, including distress and disclosure, were minimized through counseling, anonymization, secure data storage, and referral of positive cases to KCMC Hospital. The small pilot sample limits generalizability; however, it was appropriate for feasibility testing, with plans for larger validation studies.

## Implications and Recommendations

The findings of this study have important implications for autism identification and service delivery in Tanzania and similar low-resource settings. The persistence of diagnostic delays, limited availability of culturally validated screening tools, and reliance on non-standardized assessment practices highlight critical gaps in early detection systems. The acceptable reliability of the adapted M-CHAT-R/F and SCQ suggests that, when culturally tailored, these tools can be effectively integrated into community and primary care settings to support early identification of ASD. Furthermore, the influence of cultural beliefs, stigma, and reliance on traditional healers underscores the need for culturally sensitive approaches that engage both biomedical and community-based systems. The observed variability in training and awareness among stakeholders also indicates a need to strengthen workforce capacity, particularly among teachers and caregivers, who are often the first points of contact for children with developmental concerns. Additionally, the demonstrated importance of supportive communication and trust-building highlights the role of interpersonal dynamics in enhancing screening acceptability and uptake.

Based on these findings, several recommendations emerge. First, the Ministry of Education and Ministry of Health need to integrate these tools into routine early childhood development services at the community, school, and primary healthcare levels to promote early detection of ASD. Second, the Ministry of Health, in collaboration with the President’s Office–Regional Administration and Local Government (PO-RALG), teacher training institutions, and local government authorities, should develop and implement targeted training programs for non-specialist providers, including teachers, community health workers, and caregivers, to improve screening accuracy and referral pathways. Third, public awareness campaigns should be developed to address stigma and misconceptions about autism, incorporating culturally appropriate messaging and engagement with community leaders and traditional healers. Finally, to enhance feasibility among the users, screening tools should be streamlined and accompanied by clear administration guidelines to reduce time burden while maintaining diagnostic utility. Together, these strategies can contribute to more equitable, timely, and effective autism care in Tanzania.

## Conclusion

The findings provide preliminary support for the cultural appropriateness and internal consistency of the adapted tools and justify larger-scale validation studies in Tanzania. The adapted M-CHAT-R/F and SCQ showed acceptable reliability, suggesting that they can support early identification of ASD when tailored to local language, culture, and everyday experiences. The findings also revealed ongoing challenges, including stigma, limited awareness, and shortages of trained professionals, which continue to delay diagnosis and access to care. Addressing these barriers through community education, workforce training, culturally sensitive screening approaches, and stronger health and education systems could improve early detection and intervention for children with ASD in low-resource settings such as Tanzania. Future research should focus on conducting larger-scale validation studies of the culturally adapted M-CHAT-R/F and SCQ across diverse regions and populations in Tanzania to further assess their sensitivity, specificity, and overall diagnostic accuracy. In addition, future studies should explore strategies for integrating autism screening into routine community, school, and primary healthcare services, while assessing the effectiveness of training programs for teachers, caregivers, and community health workers. Research examining culturally informed awareness interventions, stigma reduction approaches, and collaboration with traditional healers may also provide valuable insights into improving acceptance, early identification, and access to autism services in low-resource settings.

## Data Availability

All data produced in the present study are available upon reasonable request to the authors

## Acknowledgement

The authors gratefully acknowledge the support provided for this study by Education Sub-Saharan Africa (ESSA) and the Research for Equitable Access and Learning Centre (REAL Centre) through Grant reference ECDAFRICA001. This project is funded by ESSA and the REAL Centre with financial support from the Conrad N. Hilton Foundation and the Global Partnership for Education Knowledge and Innovation Exchange (GPE KIX). The authors also extend their sincere appreciation to the study participants, experts, and stakeholders who contributed their time and valuable insights during the pilot implementation and adaptation process.

## Author contributions

All authors contributed to the conception and design of the study. Material preparation and data collection were performed by all authors. Data analysis was conducted by Alex G. Alexander and Aloyce Mpepo. The first draft of the manuscript was written by Alex G. Alexander, and all authors reviewed and commented on previous versions of the manuscript. Florida Joseph Muro and Blandina Mmbaga provided supervision throughout the study. All authors read and approved the final manuscript.

